# Object and Instance Detection Within Image Scenes

**DOI:** 10.1101/2021.07.03.21258850

**Authors:** Daniel Adenisimi

## Abstract

This paper compares state-of-the-art methods in object and instance detection, and examines why YOLO (You Only Look Once) outperforms top detection methods. Different Pascal VOC dataset is used as the benchmark to explore mean average precision(mAP). YOLO is twice as accurate to prior works on real-time detection. The outcome of of merging YOLO with Fast R-CNN is an increased mean average precision (mAP) which results in performance boost. Hence, YOLO is an enhanced model of top detection methods.

## 1. Introduction

Humans glance at an image and instantly know what objects are there, where they are and how they interact. The human visual system is fast and accurate, allowing the performance of complex tasks like driving with little conscious thought.

Fast and accurate algorithms for object detection would allow computers to drive cars, enable assistive devices to convey real-time scene information to humans, and unlock the potential for general purpose robotic systems.[1]

Historically, deep learning has improved image classification and object detection accuracy. But compared to image classification, object and instance detection remain a complex and challenging task. According to Redmon et. al, the complexity arises because detection requires an accurate localization of objects, which creates two primary challenges. First, candidate object locations (often called “proposals”) must be processed. Second, these proposals provide only a rough localization that must be refined to achieve precise localization. While there have been successful advances like region proposal methods and region-based convolutional neural networks (R-CNN).[2] These methods compromise on speed, accuracy, or simplicity [3]. Unlike image classification, detection requires localizing objects within an image. Hence, different scholarly articles has framed object detection as a regression problem [[3], [1]]. That is, straight from image pixels to bounding box coordinates and class probabilities. While this is the case, a new approach that asserts you only look once (YOLO) at an image to predict what objects are present and where they are has shown to be promising [1]. YOLO is extremely fast, it reasons globally about an image when making predictions and learns generalizable representations of objects[1]. This paper aims to compare state-of-the-art object and instance detection methods. Specifically, to analyze why YOLO outperforms top detection methods like R-CNN by a wide margin[1].

## 2. Related Work

This section reviews existing object and instance detection methods most related to this work.

### 2.1 Region CNN

Region-based Convolutional Network method (R-CNN) achieves excellent object detection accuracy by using a deep ConvNet to classify proposals. This detection system consist of three modules (Fig.2). The first produces category-independent region proposals (Examples include: objectness, selective search, category-independent object proposals, constrained parametric min-cuts (CPMC) and multi-scale combinatorial grouping), the second, a large convolutional neural network that extracts a fixed-length feature vector from each region, while the third is a set of class specific linear Support Vector Machines which classifies these regions.[3]

**Fig. 1:**
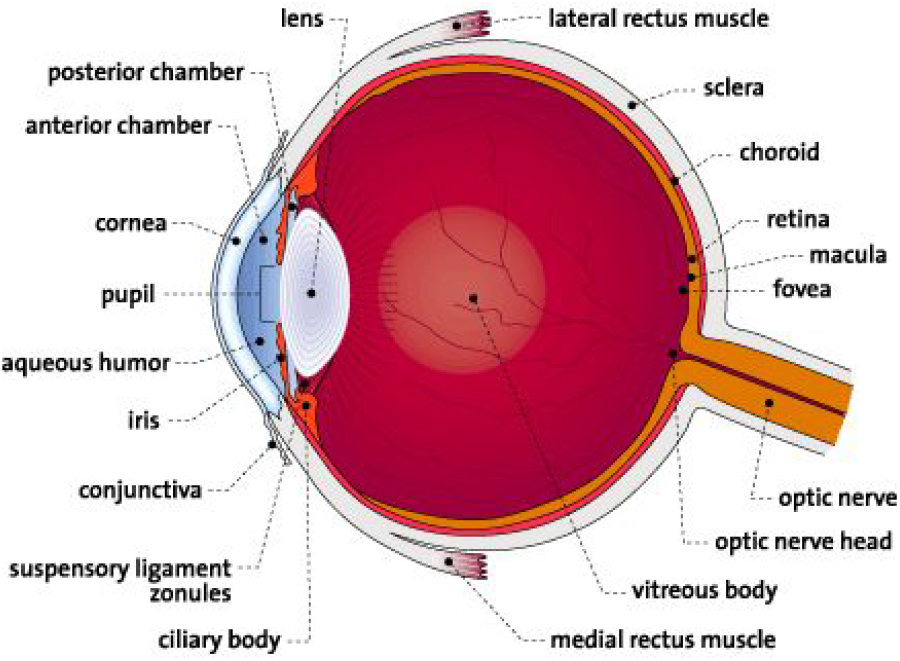
Human Eye.

**Fig. 2:**
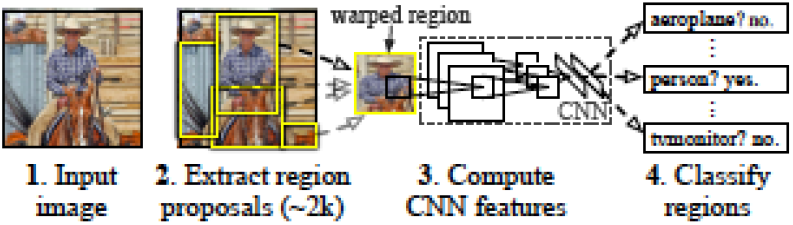
Object detection system overview. The system (1) takes an input image, (2) extracts around 2000 bottom-up region proposals, (3) computes features for each proposal using a large convolutional neural network (CNN), and then (4) classifies each region using class-specific linear SVMs.[3]

**Fig. 3:**
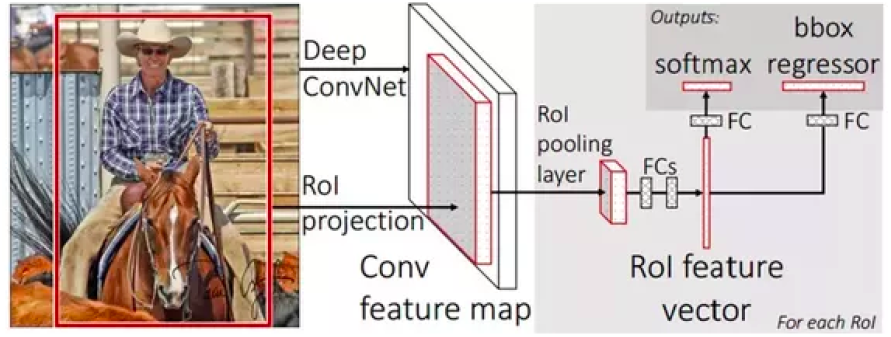
Fast R-CNN architecture. An input image and multiple regions of interest (RoIs) are input into a fully convolutional network. Each RoI is pooled into a fixed-size feature map and then mapped to a feature vector by fully connected layers (FCs). The network has two output vectors per RoI: softmax probabilities and per-class bounding-box regression offsets.[3]

**Fig. 4:**
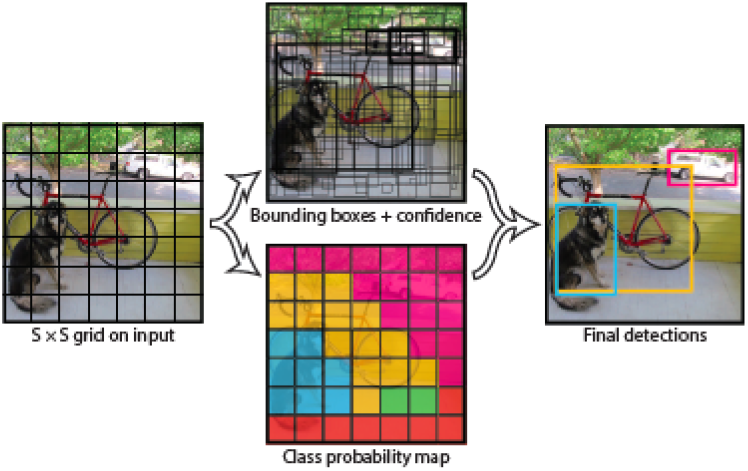
The Model. Image is divided into an S x S grid. For each grid cell, it predicts B bounding boxes, confidence for those boxes and C class probabilities.[1]

**Fig. 5:**
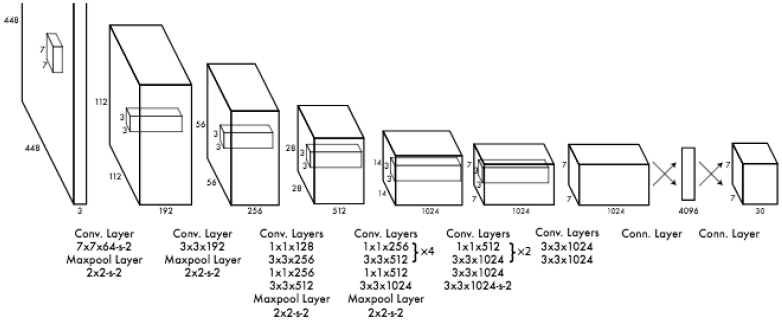
The Architecture. Yolo detection network has 24 convolutional layers followed by 2 fully connected layers.[1]

Although R-CNN achieves excellent detection accuracy, it has setbacks that includes:

1. **Multi-stage training pipeline**: R-CNN must first fine-tune a ConvNet on object proposals using log loss. It furthers needs to fit an algorithm that sorts data into two categories called support vector machines (SVMs) to ConvNet features. Where SVMs act as object detectors replacing the softmax classifier learnt by fine-tuning. In the third training stage, bounding-box regressors are learned. [3]
2. **Expensive training in space and time**: To train SVMs and bounding-box regressors, features are extracted from each object proposal in each image and written to disk. For example, in a deep network architecture like the Oxford trained Visual Geometry Group 16 (VGG16), the process takes 2.5 GPU-days for the 5k resolution images of the VOC07 trainval dataset. And the features also requires hundreds of gigabytes of storage. [3]
3. **Slow Object Detection**: At test-time, features are extracted from each object proposal in each test image. For instance, VGG16 detection takes 47s / image on a graphics processing unit (GPU). [3]

To sum up, R-CNN is slow because it performs a ConvNet forward pass for each object proposal, without sharing computation. [3]

### 2.2 Fast Region CNN

Fast R-CNN takes as input an entire image and a set of object proposals. First, the network processes the whole image with several convolutions and max-pooling layers to produce a convolutional feature map. Then, for each object proposal, a region of interest (ROI) pooling layer extracts a fixed-length feature vector from the feature map. Each feature vector is then fed into a sequence of fully connected (fc) layers that finally branch into two sibling output layers: one that produces softmax probability estimates over K object classes plus a catch-all “background” class and another layer that outputs four real-valued numbers for each of the K object classes which defines the bounding-box positions for one of the K classes. Therefore, it can be noted that Fast R-CNN method has several advantages over R-CNN, such as:

1. Higher detection quality mean average precision (mAP) than R-CNN
2. Training is single-stage, using a multi-task loss
3. Training can update all network layers
4. No disk storage is required for feature caching [3]

### 2.3 Faster Region CNN

Faster R-CNN is similar to Fast R-CNN, but applies Region Proposal Networks (RPNs) that shares convolutional layers with state-of-the-art object detection networks. That is, instead of using Selective Search (SS) which greedily merges superpixels based on engineered low-level features and is an order of magnitude slower (2s per image) in a CPU implementation when compared to efficient detection networks, RPNs are trained end-to-end specifically, for the task of generating detection proposals. The training scheme alternates between fine-tuning for the region proposal task and then fine-tuning for object detection, while keeping the proposals fixed. The scheme converges quickly and produces a unified network with convolutional features that are shared between both tasks. This scheme doesn’t only rely on inexpensive features but also enables nearly cost-free region proposals given the detection network’s detection. And with a simple alternating optimization, RPN and Fast R-CNN can be trained to share convolutional features. [4]

## 3. You Only Look Once (YOLO)

YOLO is a unified system that is able to detect the potential region of interests (ROIs) from an entire whole image and directly predict their class probabilities[5]. It posses a single neural network that uses features from an entire image to predict bounding boxes across all classes of the image. [1]

YOLO does this by taking an input image and dividing it into an S x S grid. Each grid cell then predicts bounding boxes and confidence scores, including class probabilities for those boxes. The bounding box each consists of 5 predictions: x, y, w, h, and confidence. The (x, y) coordinates represent the center of the box in relation to the bounds of the grid cell. The width and height (w, h) are predicted relative to the whole image. In the same way, the confidence prediction represents the intersection over union (IOU) between the predicted box and any other ground truth box. [1]

During test time, the conditional class probabilities and the individual box confidence predictions are multiplied which then gives a class-specific confidence scores for each box. These scores encode both the probability of that class appearing in the box and how well the predicted box fits the object. [1]

Furthermore, YOLO-based deep learning technique has been employed in different fields. Examples include: a. Computer aided diagnoses (CAD) in the detection of breast cancer where a trained YOLO-based CAD system detects cancer masses and classifies them into benign or malignant[5]. b. Vehicle detection, where it achieved comparable results with state-of-the-art methods on several data sets, while being significantly faster[6].YOLO’s application in breast cancer detection is based on the findings of M. A. Al-masni et al. proposal of four main stages: mammogram preprocessing, feature extraction utilizing multiconvolutional deep layers, mass detection with confidence model, and breast mass classification using fully connected neural network (FC- NN). In fact, the results of the proposed CAD system show its ability to detect the location of benign and malignant masses as illustrated in Fig. 6. When compared to the ground truth in Fig. 6(a) and (c), YOLO can exactly detect the masses as shown in Fig.6(b) and (d). At the time, YOLO’s abnormalities detection performance of benign and malignancy of the breast in mammograms was shown to have an overall accuracy of 96.33%.[5].

**Fig. 6:**
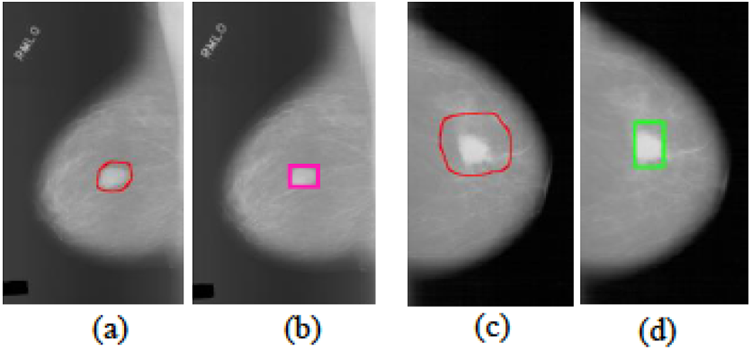
Mass detection. (a) and (b) show the ground-truth mass over the pectoral muscle and the detected by Al-masni et al. proposed method, respectively. (c) and (d) present the ground-truth mass surrounding by dense tissue and the detected by Al masni et al. proposed method, respectively.[5]

## 4. Experiment

To prove the hypothesis that YOLO outperforms top detection methods, YOLO is first compared with deformable parts model (DPM) and then with other real time detection variants like R-CNN [1]. This comparison is based on different Pascal Visual Object Classes (VOC) data sets, a publicly available dataset of images and annotation together with a standardised evaluation procedure [7]. Furthermore, the methodology and tools of Hoiem et al. is used to compare errors made by YOLO and Fast R-CNN (one of the highest performing versions of R-CNN) on VOC 2007 [1]. Finally, VOC 2012 and 2007 is used as the benchmark for comparing YOLO’s performance when it is combined with R-CNN and the result of it’s mean average precision (mAP) to current state-of-the-art methods.[1]

## 5. Results

Fast YOLO is the fastest object detection method on PASCAL when compared with the GPU implementation of DPM which runs at 30Hz or 100Hz. With 52.7% mAP, Fast YOLO is twice as accurate to prior work on real-time detection and it retains real-time performance when it’s mAP is pushed to 63.4%. [1]

On VOC 2007, Fast R-CNN achieves mAP 66.0% [3] and helps speed up the classification stage of R-CNN [1]. Nevertheless, selective search remains its requisite which takes around 2 seconds per image to generate bounding box proposals. Therefore, regardless of Fast R-CNN high mAP, at 0.5 fps, it is still far from realtime detection when compared to YOLO. [1] While this is the case, YOLO still struggles to localize objects correctly leading to a lot of localisation errors[Fig. 7]. Whereas Fast R-CNN, which is also prone to background errors is still 3 times more likely to predict background detections than YOLO. [1]

**Fig. 7:**
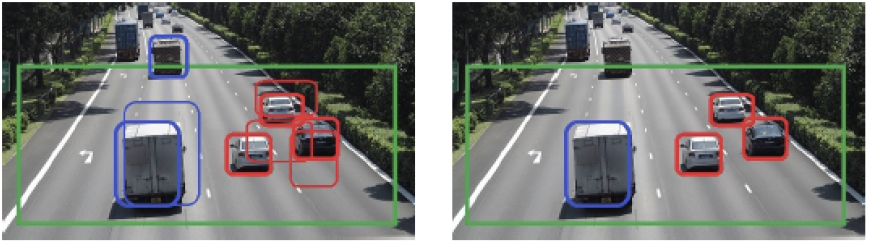
Examples for vehicle detection approach on a road image. The green rectangle is the selected road region for detection. Red and blue rectangles in (a) are the initial detection results by YOLO model. After removing invalid detection results, the final detection results are shown in (b).[6]

**Fig. 8:**
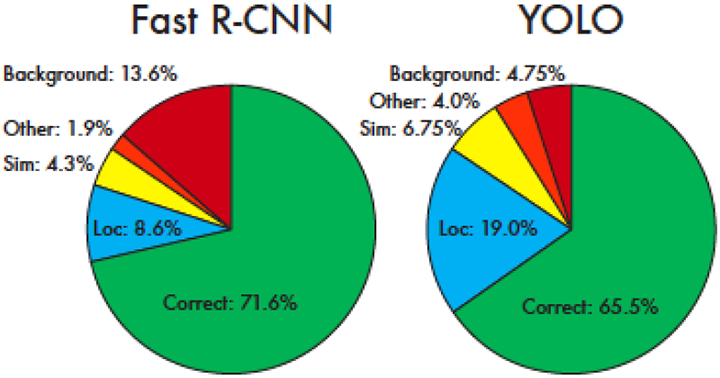
Error Analysis: Fast R-CNN vs. YOLO. These charts show the percentage of localization and background errors in the top N detections for various categories (N = # objects in that category)[1]

However, when YOLO is combined with Fast R-CNN, it eliminates the background detections from Fast R-CNN leading to a significant boost in performance and making it one of the highest performing detection methods on the public leaderboard on VOC 2012 test. In short, when two separate Fast R-CNN with mAPs 71.8% and 68.4% on VOC 2007 and 2012 were combined with YOLO. Their mAPs increased significantly, at 75% and 70.7% respectively. [1]

## 6. Discussion and Conclusion

This paper proposes YOLO is simple and straight forward for object and instance detection when compared to state-of-the-art methods.YOLO outperforms other detection methods by quickly predicting potential region of interest and class probabilities of an entire image. [1] The result of Fast YOLO’s fps (155) and YOLO’s stand-alone mAP of 63.4% and 75% if combined with Fast R-CNN on Pascal VOC 2007 is a testament that YOLO is faster and twice as accurate. Therefore, YOLO is an optimised model of other R-CNN variants and further research can be done to explore models that reduces YOLO’s localization errors. [1]

## Data Availability

Publicly available

